# Blink rate measured *in situ* decreases while reading from printed text or digital devices, regardless of task duration, complexity or working distance

**DOI:** 10.1101/2022.11.19.22282503

**Authors:** Ngozi Charity Chidi-Egboka, Isabelle Jalbert, Jiaying Chen, Nancy E. Briggs, Blanka Golebiowski

**Affiliations:** School of Optometry and Vision Science, Faculty of Medicine and Health, UNSW Sydney, NSW 2052, Australia; Mark Wainwright Analytical Centre, UNSW Sydney

**Author notes:** **Corresponding author:** Ngozi C. ChidiEgboka, School of Optometry and Vision Science, Faculty of Medicine and Health, Level 3, North Wing, Rupert Myers Building, Gate 14 Barker St, UNSW Sydney, NSW 2052, Australia. **Commercial relationship disclosures 1.** Ngozi Charity Chidi-Egboka, None declared; **2.** Isabelle Jalbert, None declared; **3.** Jiaying Chen, None declared; **4.** Nancy E. Briggs, None declared; **5.** Blanka Golebiowski, None declared.

**Keywords:** Blinking, Dry eye, Smartphone, Ocular Surface, Reading, Repeatability, Digital device

## Abstract

**Purpose:** To compare blinking measured *in situ* during various tasks and examine relationships with ocular surface symptoms. Day-to-day repeatability of blink rate and interblink interval was assessed.

**Methods:** Twenty-four students (28.6±6.3 years; 8M:16F) completed six reading tasks (printed text, laptop, TV, smartphone, smartphone at 50% brightness, smartphone with complex text), and two non-reading tasks (conversation, walking) in a randomised cross-over study. Ocular surface symptoms and clinical signs were assessed. Blink rate and interblink interval were measured using a wearable eye tracking headset. Blink parameters were compared across tasks and time (linear mixed model and post hoc comparisons with Bonferroni correction). Associations between blinking, symptoms, ocular surface, and clinical signs were assessed (Spearman’s correlation). The smartphone reading task was completed twice to determine coefficient of repeatability.

**Results:** Blink rate was lower (mean 10.7±9.7 blinks/min) and interblink interval longer (mean 9.6±8.7s) during all reading tasks compared to conversation (mean 32.4±12.4 blinks/min; 1.5±0.6s) and walking (mean 31.3±15.5 blinks/min; 1.9±1.3s) (p<0.001). There were no significant differences in blink parameters between any of the reading tasks, nor between conversation and walking. Changes in blinking occurred within one minute of starting the task. No associations were evident between blink rate or interblink interval and ocular surface symptoms or signs. Coefficient of repeatability was ±12.4 blinks/min for blink rate and ±18.8s for interblink interval.

**Conclusion:** Spontaneous blinking can be reliably measured *in situ*. Blink rate was reduced and interblink interval increased during reading compared to conversation and walking. Changes in blinking were immediate and sustained, and not associated with ocular surface symptoms or signs.

## 1. Introduction

Blinking maintains a stable tear film, thereby sustaining ocular surface integrity and visual function [1]. Disruptions to blinking disturb ocular surface homeostasis and may contribute to ocular discomfort and dry eye [2, 3].

Blinking is affected by the type, complexity, and cognitive demand of the task undertaken during measurement [4-8]. Differences in viewing distance, factors such as font size, contrast and device used create different demands on blinking [5, 9]. Previous studies have found increased discomfort linked to impaired blinking during smartphone and computer use [3, 9-11]. Blinking has been investigated during various tasks, (e.g., conversation, reading, playing computer games, watching a film, listening to music, resting quietly) of various complexities and on various devices including printed text, desktop and laptop computer, tablet, and, at various viewing distances and gaze positions [11-18]. However, blink assessment remains hampered by lack of a gold standard method and standardised conditions of measurement.

A wide range of mean blink parameters have been previously reported ranging from 11 – 36 blinks/min during conversation, 4 – 14 blinks/min during reading, 5 – 26 blinks/min during rest and directed fixed gaze, in adults [19]. This wide range can be explained in part by differences in definitions of spontaneous blinking. Various definitions of a blink include a ‘25% downward movement of the upper eyelid’ from the fully open position [20], an ‘obvious downward eyelid movement’ [21], the ‘upper eyelid reaching downwards from the top of the pupil’ [22], a ‘downward movement of the upper eyelid covering 30%–75% of the cornea’ [7] and a ‘15% decrease in the height of the upper eyelid’ [23]. Blinking is difficult to assess clinically or in situ outside of the laboratory setting, thus a method that allows more natural measurement may be helpful in standardising the definition of spontaneous blinking.

Blink measurements in previous studies have typically occurred in settings not representative of real-life situations, requiring participants to keep a stationary head position on a chin and forehead rest which may limit the complete view of the anterior eye during measurement [19]. Fixed head positions during measurement may stimulate participants awareness and impact the accuracy of blink parameters [24]. Robust blink measurement requires the whole anterior eye to be constantly visible so that the full range of eyelid movements can be observed [25]. A higher than 95% blink detection accuracy in relation to pupil detection has previously been demonstrated with head mounted eye-tracking technology which allows free head position [25]. Measuring spontaneous blink activities in real time under real-life situation is desirable to improve understanding of blink behaviour and for the relationship between ocular symptoms and blink parameters to be adequately characterised.

A recent study in children demonstrated that the blinks counted by the Pupil software blink detection algorithm using the wearable eye tracker were in agreement with a manual count [26]. Hence blinking *in situ* could be reliably measured using a wearable eye tracking headset, showing a rapid decrease in blink rate during one hour of smartphone gaming, which was linked to ocular discomfort [26, 27]. However, it is not clear if this effect was due to the use of smartphones per se, or due to the task of reading itself.

The repeatability of recent and commonly used blink measurement methods has not been assessed. Repeated measurements of blink rate have been reported for electrophysiology methods (magnetic search coil technique and electro-oculography) [6, 18, 28], and for manual counting of blinks from eye video recording [29-31] conducted in a laboratory setting where participants’ head position was fixed. However, none of these studies reported standard measures of repeatability [32].

The current study aimed to compare blink parameters (blink rate, interblink interval) during various reading and non-reading tasks measured *in situ* using a wearable eye tracking headset and to examine associations with ocular surface symptoms. In addition, the day-to-day repeatability of blink rate and interblink interval measurement was assessed.

## 2. Methods

A randomised cross-over open label study was conducted. Approval was obtained from the UNSW Human Research Ethics Advisory Panel and the tenets of the Declaration of Helsinki were adhered to. Informed consent was obtained from all participants prior to participation.

### 2.1 Participants

Students aged 18-40 years were recruited from the UNSW Sydney campus. Minimum unaided visual acuity of 0.1LogMAR at 6m and 40cm and binocular vision (accommodation and convergence) normal for age were required for participants to be enrolled in the study including a minimum amplitude of accommodation of 5D (push up to blur with Royal Air Force rule (RAF rule)) and a near phoria equal or smaller than 6 prism dioptres (modified Thorington test) [33]. Participants were excluded if they wore spectacle or contact lenses or had a history of ocular conditions including eye allergies, systemic conditions (e.g., Parkinson’s disease, diabetes) or medications (e.g., cornea cold thermoreceptor stimulants such as menthol ointment; dopamine antagonist drugs) likely to impact blinking [34, 35]. Sample size calculation (SAS 9.4 (2012) NC, USA) showed that 24 participants were required to detect a difference in blink rate between various tasks of 5.8 blinks/min [12, 15], with 90% power at alpha (α) level of 0.05/7 (statistical significance corrected for multiple comparison of seven conditions) and to account for a possible 20% attrition. Twenty-four participants were also sufficient to assess day-to-day repeatability of blink rate and interblink interval, based on a desired precision of ±30% which was expressed as a percentage of within-person standard deviation, with two repeated measurements.

### 2.2 Procedures

All participants attended two visits (Figure 1) during which they completed a questionnaire on demographics and daily hours of digital device use, and eight tasks as described below. Ocular symptoms and the ocular surface were assessed and *in situ* blinking was measured. In line with the COVID-19 safety protocol and guidelines which came into effect in Sydney, Australia, part-way through the study, some of the participants wore a surgical mask that covered from nose to chin for all assessments during both study visits (Figure 2c, 2d).

**Figure 1.**
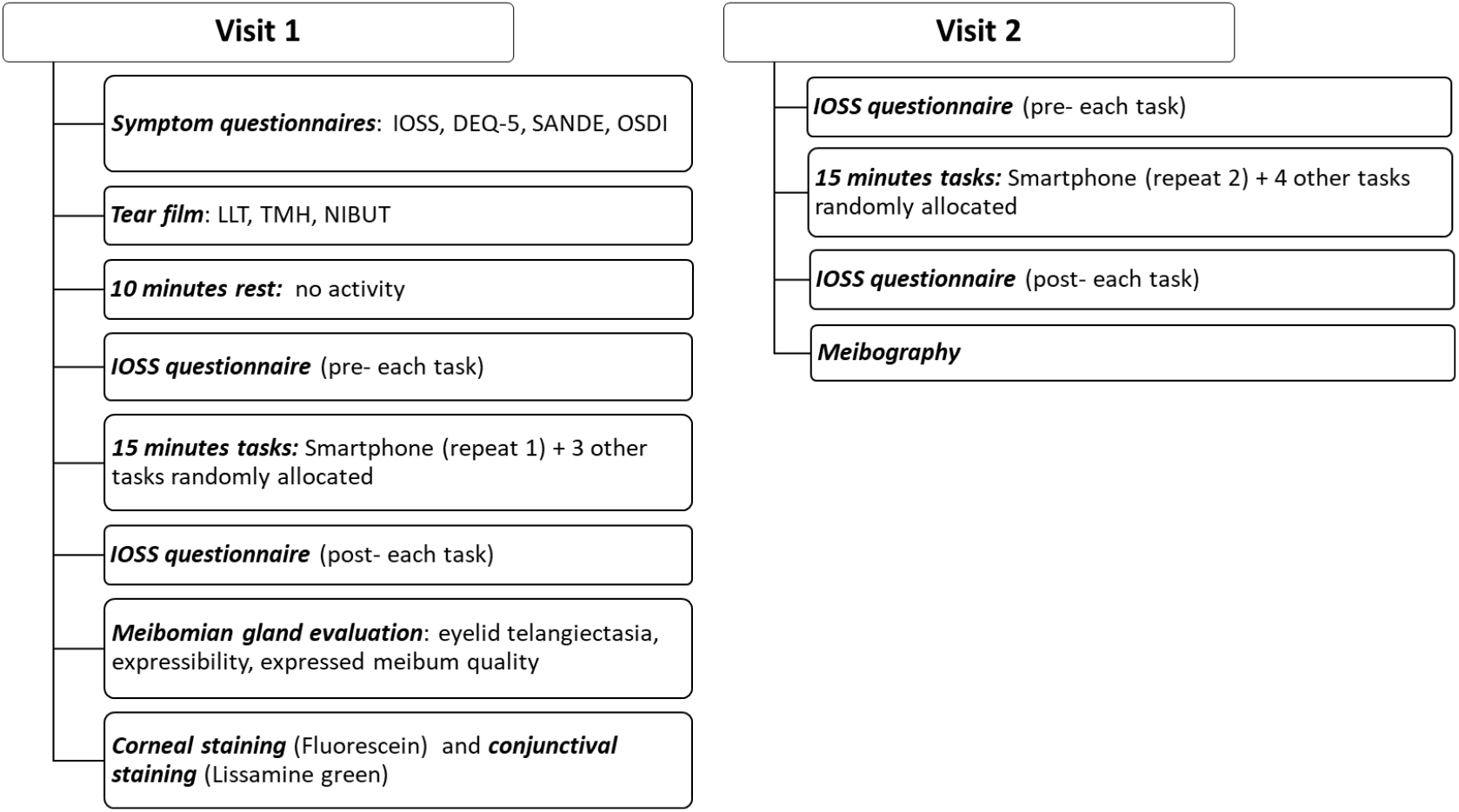
Flowchart of study visits and order of clinical assessments. Note: Visit 2 was conducted two days after visit 1. Smartphone task was completed twice before other tasks at each visit for assessment of repeatability. Other tasks randomly allocated include: six reading tasks (printed text, laptop, smart TV at 6m, smartphone, smartphone at 50% brightness, smartphone more complex text), and two non-reading tasks (conversation, walking indoors). IOSS - Instant Ocular Symptoms Survey; DEQ-5 - Dry Eye Questionnaire 5; SANDE - Symptoms Assessment in Dry Eye; OSDI - Ocular Surface Disease Index; LLT - Lipid layer thickness; TMH - Tear meniscus height; NIBUT - Non-invasive tear break-up time.

**Figure 2.**
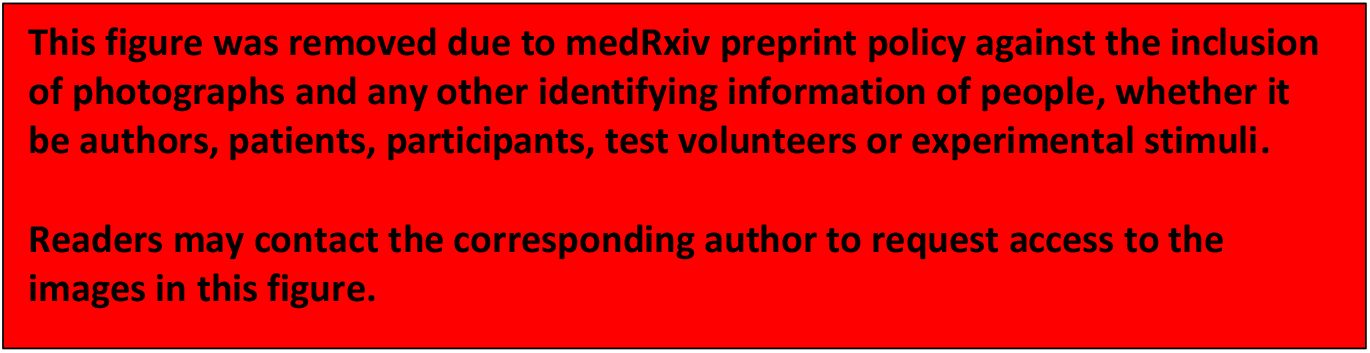
Study set-up showing the wearable eye tracking headset (Pupil Labs GmbH Berlin, Germany) with two inbuilt high-speed eye cameras and a scene camera for real time monitoring from participants’ vantage point. The headset was worn by study participants during various tasks including reading from **a)** printed text, **b)** laptop, **c)** smart TV at 6m, **d)** smartphone, **e)** walking indoors and conversation (not shown). The wearable eye tracking headset was connected to a laptop for task monitoring and data acquisition for all tasks other than walking indoors, where an android phone was used for the same purpose, while the examiner followed behind the participant holding the android phone to monitor recording (Figure 2e). Participants’ consents were obtained for use of these images.

#### 2.2.1 Tasks

The tasks comprised of six reading tasks (printed text, laptop, smart TV at 6m, smartphone, smartphone at 50% brightness, smartphone with more complex text), and two non-reading tasks (conversation, walking indoors). All tasks were of 15 minutes duration. Tasks were completed in random order, other than the smartphone task, which was completed first at each visit (repeated twice) (Figure 1). Data for the repeat 1 of the smartphone task was used for all analyses except for repeatability where both repeats were used. A break of approximately three minutes was allowed between tasks to allow completion of a pre- and post-task questionnaire (section 2.2.2).

A reading level 5th to 7th grade was selected for the reading task [36, 37]. A text of reading level of university graduate was selected for the complex smartphone reading task [38]. For all reading tasks, the default text font size was 16 pixels (equivalence of 12 points), black Times New Roman. However, the viewing distances, screen or display size and the varying digital device pixel may affect the actual angular extent and therefore alter the font sizes [39, 40]. The printed text reading task was printed one-sided in A4 format. Conversation was elicited using age-appropriate ‘great conversation starters’ [41]. The walking indoors task was conducted in a level corridor of a temperature-controlled university building.

The same smartphone (iPhone 8 Plus, 5.5-inch, 1920 × 1080 pixel at 401ppi, 2017) was used for all smartphone reading tasks. A MacBook Pro (13.3-inch, 2560×1600 ppi built-in display, 2019) was used for the laptop task, and a smart TV (NEC, Model: V754Q, 75-inch, 3840×2160 ppi) for reading at 6m (Figure 2). Participants were instructed to hold the smartphone at their habitual reading distance, to use one finger to scroll to the next page or the side arrow button on the laptop keyboard to scroll to the next page during reading, and to not alter the screen brightness or font size. The smartphone was set at maximum screen brightness of measured luminance 380 cd/m^2^ (Konica Minolta CS-100A) for two tasks and reduced to half during from the smartphone at 50% brightness task (measured luminance 121 cd/m^2^). The laptop screen and smart TV were also set at maximum screen brightness, measured luminance 316 cd/m^2^and 316 cd/m^2^ respectively. The measured luminance for the printed text reading was 77 cd/m^2^.

#### 2.2.2 Ocular symptoms and ocular surface clinical assessments

Baseline ocular surface symptoms were assessed using the Instant Ocular Symptoms Survey (IOSS) [42], Dry Eye Questionnaire 5 (DEQ-5) [43], Symptoms Assessment in Dry Eye (SANDE) [44], and Ocular Surface Disease Index (OSDI) [45] questionnaires, self-completed by participants. The IOSS questionnaire (printed text) was completed by participants between (pre- and post-) tasks. The IOSS was found to be an effective tool for instant symptom measurement, with good diagnostic ability and was developed to measure instantaneous symptoms, i.e., at the time of administration (compared to the other questionnaires which record symptoms experienced over the preceding weeks), and as such is appropriate to administer for repeated comfort assessment [42].

The following baseline tear film clinical assessments were conducted prior to blink measurements: tear film lipid layer thickness (LLT) (LipiView® interferometer; Tear Science, Morrisville, NC), tear meniscus height (TMH), and non-invasive tear break-up time (NIBUT), (Oculus® Keratograph 5; Oculus®, Arlington, WA). The index of LLT based on mean interferometry colour units was recorded [46]. TMH was assessed in the regions vertically below the pupil centre, and directly under the nasal and temporal corneal limbal edge (determined using the integrated ruler) to account for variability in TMH along the length of the lower meniscus, and the average of the three measurements was recorded [47]. The automated detection of the first tear break-up was recorded for NIBUT [48]. Measurement of the tear breakup time with NIBUT technique was considered preferable [49, 50], because it is automated compared to other subjective methods e.g., videokeratoscope, Tearscope, with which measurements have been found to vary between sessions and observers [50, 51].

Ocular surface clinical assessments were performed on the right eye only, in the same temperature-controlled examination room, in ascending order of invasiveness [50]. General ocular surface health, corneal staining (Fluorescein) and conjunctival staining (Lissamine green strips, GreenGlo™) (Oxford grading scale) [52, 53], telangiectasia [54], meibomian gland expressibility, meibography imaging of the upper eyelid (Oculus® Keratograph 5; Oculus®, Arlington, WA) was scored in relation to loss of meibomian glands using the meiboscore (meiboscore: degree 0=no gland loss, 1≤25% gland area of loss, 2=26%–50% gland area loss, 3=51%–75% gland area loss, 4≥75% gland area loss) [55] and the pattern of meibomian gland morphological changes were assessed [55-57] after all tasks were completed as shown in Figure 1.

#### 2.2.3 *In situ* Blink measurement

Blink assessment was conducted after tear film assessment, following 10 minutes of rest. *In situ* assessment of blink parameters was conducted during each task using a binocular wearable eye tracking headset (Pupil Labs Core GmbH Berlin, Germany) [58] (Figure 2). Data were analysed using mean values for each minute as well mean values over 12 or 15 minutes of recording.

The wearable eye tracking headset recorded participants’ eyes using the two inbuilt eye cameras with a resolution of 192×192 pixels at 120 Hz (Figure 2) [58]. The eye camera (providing a view of the participant and their eye) together with the scene camera (providing a view of what the participant is looking at) (Figure 2) enabled continuous monitoring of participant adherence in real-time. Blink activity was detected using the open-source eye tracking software Pupil v2.0 (Pupil Labs GmbH Berlin, Germany), based on visibility of the pupil as previously described [26, 58]. Briefly, the Pupil software assigns a quality measure for the detected pupil in each video frame, referred to as ‘pupil confidence’. The pupil confidence value indicates how accurately the edge of the detected pupil fits an ellipse (range: 0 (no fit) to 1 (good fit) [26, 58]. Blinks are assumed to occur during pupil confidence drops evident when the pupil is obscured, hence pupil confidence is a proxy measure for blink detection [58]. Blink data was extracted from the eye tracker recordings using Pupil software Player module (Pupil Labs GmbH Berlin, Germany) as CSV files [26]. Blink rate (number of blinks per minute) and interblink interval (the time between the end of one blink to the start of the following blink) data were estimated using Pupil software blink detection algorithm as described elsewhere [26].

For the reading from a smartphone task (repeats 1 and 2), data from the first three minutes of video recording were discarded and the remaining 12 minutes used for analysis, to allow for adjustment and adaptation to wearing the headset as recommended [59]. Complete recordings (15 minutes) were analysed for all other tasks, as participants continued with each subsequent tasks without removing the headset.

### 2.3 Repeatability of blink measurements

Participants completed the reading from smartphone task with maximum screen brightness twice at separate study visits occurring two days apart at the same time of day (between 10 am and 11 am). Time of the day was controlled as blink rate has been reported to exhibit diurnal variation (higher in the evening) [60].

### 2.4 Statistical analysis

Statistical analysis was performed using IBM SPSS Statistics (version 26, 2019; Armonk, NY, USA). A linear mixed model with fixed effect of task and mask wear and their interactions was used to examine differences in blink parameters between tasks and the effect of mask wearing on the differences in blink parameters. A separate linear mixed model with fixed effect of time was used to compare differences in blink parameters across time, within each task. Another model with fixed effect of task, time and mask wear was used to examine the differences in ocular symptoms across tasks and time. All models included a random effect for individual to account for repeated measures within-person. Model-estimated means were obtained and post-hoc pairwise comparisons were performed between tasks, between each minute within each task duration, and between pre- and post-task symptoms within each task, and p-values corrected for multiple comparisons by a Bonferroni adjustment. Spearman’s bivariate correlation was used to examine associations between blinking and changes in ocular symptoms, and ocular surface and tear film indices; p-values for the correlations were adjusted for multiple comparisons using the one-step Bonferroni method. The statistical approach suggested by Bland and Altman was used to examine repeatability of blink rate and interblink interval. The coefficient of repeatability (CoR = 1.96 x SD of differences between the two repeats), mean difference (bias) between repeats and limits of agreement (LOA = bias±CoR) were calculated and paired t-tests were used to examine agreement between repeats [32]. Significance was established at p ≤ 0.05.

## 3. Results

Twenty-four participants with normal ocular surface health completed the study. Participants were aged 18 to 40 years (mean 28.6±6.3 years), 67% were female and comprised different ethnicities: African (38%), South Asian (21%), Middle Eastern (17%), East Asian (12%), Caucasian (12%). Fourteen participants wore a surgical mask during data collection.

Thirteen data points were excluded, where more than 60% pupil confidence values were below 0.6, as per the manufacturer’s recommendation [26]: three from the printed text task, one from smart TV, five from smartphone, two from smartphone (50% brightness), and two from smartphone (more complex text). Ocular surface symptoms and clinical signs reported by participants who had excluded data points were within the range of other participants.

Baseline ocular surface symptoms and clinical assessments are presented in Table 1. Examination room temperature was maintained at 21.9±0.7°C.

**Table 1:** Baseline ocular surface symptoms and clinical assessments for 24 students with healthy eyes. Data are presented as mean±SD (range) and median (range). Higher eye symptom questionnaire scores indicate worse discomfort. IOSS - Instant Ocular Symptoms Survey; DEQ-5 - Dry Eye Questionnaire 5; SANDE - Symptoms Assessment in Dry Eye; OSDI - Ocular Surface Disease Index.

| Variables | Value |
| --- | --- |
| **Ocular surface symptoms** (score) |  |
| IOSS (0 – 10) | 1.9±1.6 (0 – 5) |
| DEQ-5 (0 – 22) | 5.9±4.1 (0 – 16) |
| SANDE (0 – 100) | 18.1±22.9 (0 – 81) |
| OSDI (0 – 100) | 15.1±16.6 (0 – 61) |
| **Tear film** |  |
| Lipid layer thickness (nm)* | 43.0±18.8 (20 – 75) |
| Tear meniscus height (mm) | 0.29±0.08 (0.17 – 0.47) |
| Non-invasive tear break-up time (s) | 10.3±6.7 (3.4 – 22.9) |
| **Cornea white light staining** (grade) | 0 (0 – 0) |
| **Meibomian gland evaluation** (grade) |  |
| Eyelid telangiectasia (grade) | 0 (0 – 2) |
| Expressibility (number of expressible glands) | 2 (0 – 3) |
| Expressibility (amount of pressure applied) | 2 (0 – 3) |
| Expressed meibum quality (grade) | 0 (0 – 1) |
| **Cornea staining - fluorescein** (0 – 15) | 0 (0 – 0) |
| **Conjunctival staining - Lissamine green** (0 – 15) | 0 (0 – 2) |
| **Meibography** |  |
| Meibomian gland area loss (score) | 1 (0 – 3) |
| Meibomian gland morphological pattern present: |  |
| Dilation | 13 (54%) |
| Shortening | 18 (75%) |
| Tortuosity | 14 (58%) |

\*Data for 21 participants only included because lipid layer thickness values measuring above the upper cut-off of 100 “interferometric colour units (ICU)” were not displayed by the instrument for three participants.

### 3.1 Differences in blink parameters between tasks

There were significant differences in blink rate (F = 29.94, p<0.001) and interblink interval (F = 38.32, p<0.001) between tasks. Blink rate was lower and interblink interval was longer during all reading tasks compared to conversation (p<0.001) and walking indoors (p<0.001) (Figure 3). There were no significant differences in blink rate or interblink interval between conversation and walking indoors, nor between any of the reading tasks. Interactions between tasks and mask wear were not significant, indicating that mask wear did not have an effect on the differences in blink rate (p=0.65) or interblink interval (p=0.72) between tasks. Blink rate and interblink interval remained unchanged throughout measurement duration for each task (p>0.05) (Figure 4).

**Figure 3.**
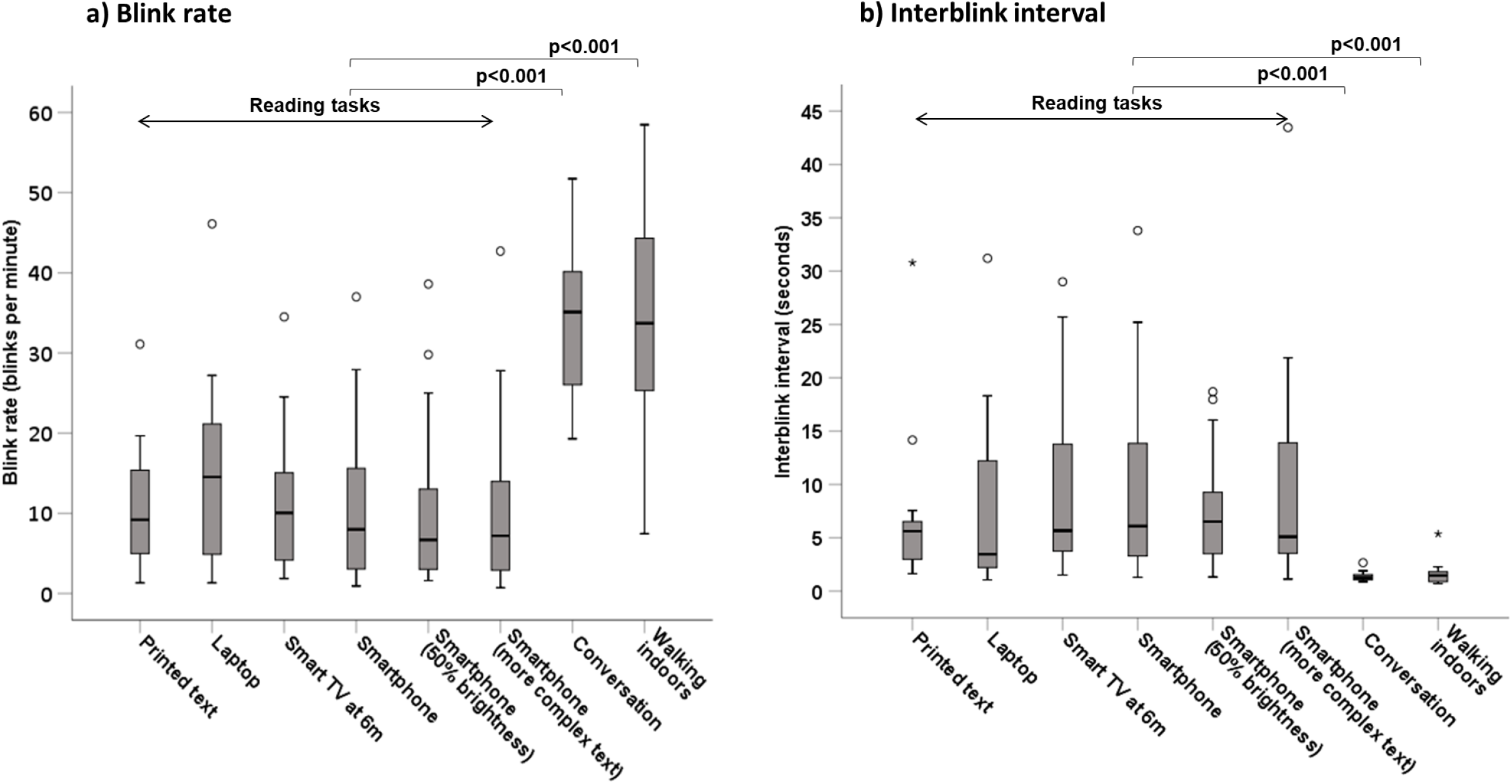
**a)** Blink rate and **b)** Interblink interval during various tasks of 15 minutes duration, measured using a wearable eye tracking headset (Pupil Labs GmbH Berlin, Germany) for 24 students with healthy eyes. Note, data from the first three minutes of the smartphone task were discarded and the remaining 12 minutes used for analysis. Data are presented as median and interquartile range. *Open circles represent mild outliers (measurements >1*.*5 to 3 times the interquartile range) and stars represent extreme outliers (measurements >3 times the interquartile range)*.

**Figure 4.**
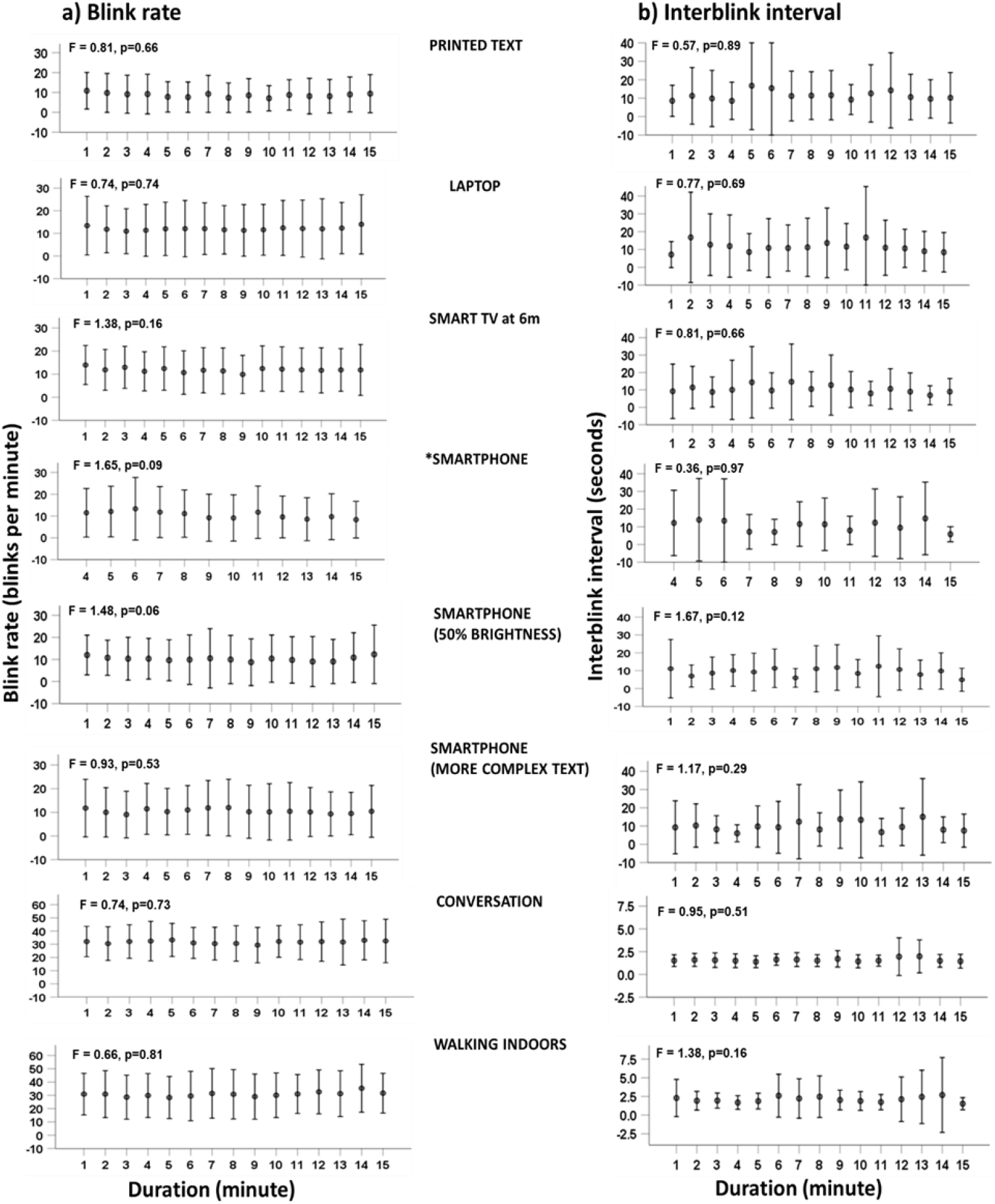
**a)** Blink rate and **b)** Interblink interval during various tasks of 15 minutes duration, measured using a wearable eye tracking headset (Pupil Labs GmbH Berlin, Germany) for 24 students with healthy eyes. The tasks include: six reading tasks (printed text, laptop, smart TV at 6m, smartphone, smartphone at 50% brightness, smartphone more complex text), and two non-reading tasks (conversation, walking indoors). *Note, data from the first three minutes of the smartphone task were discarded and the remaining 12 minutes used for analysis.

### 3.2 Differences in ocular symptoms pre- and post-task and association with blinking

Ocular symptoms (IOSS) pre- and post-tasks differed between tasks (F = 4.69, p<0.001). Symptoms worsened after reading from a smartphone when text was more complex (p=0.01) or at 50% brightness (p=0.02), and from a smart TV (p<0.001) but did not change during other tasks (Figure 5). There was no evidence that mask wearing influenced these differences (mask wear*task*time interaction, p=1.00). These changes in symptoms were not associated with blink rate or interblink interval (rho -0.09 to 0.41, p=1.00) (supplementary Table 1). There were no associations between blinking and baseline ocular surface symptoms (OSDI, SANDE, DEQ-5, IOSS), tear film and other clinical indices (rho -0.01 to 0.45, p=1.00) (supplementary Table 2).

**Figure 5.**
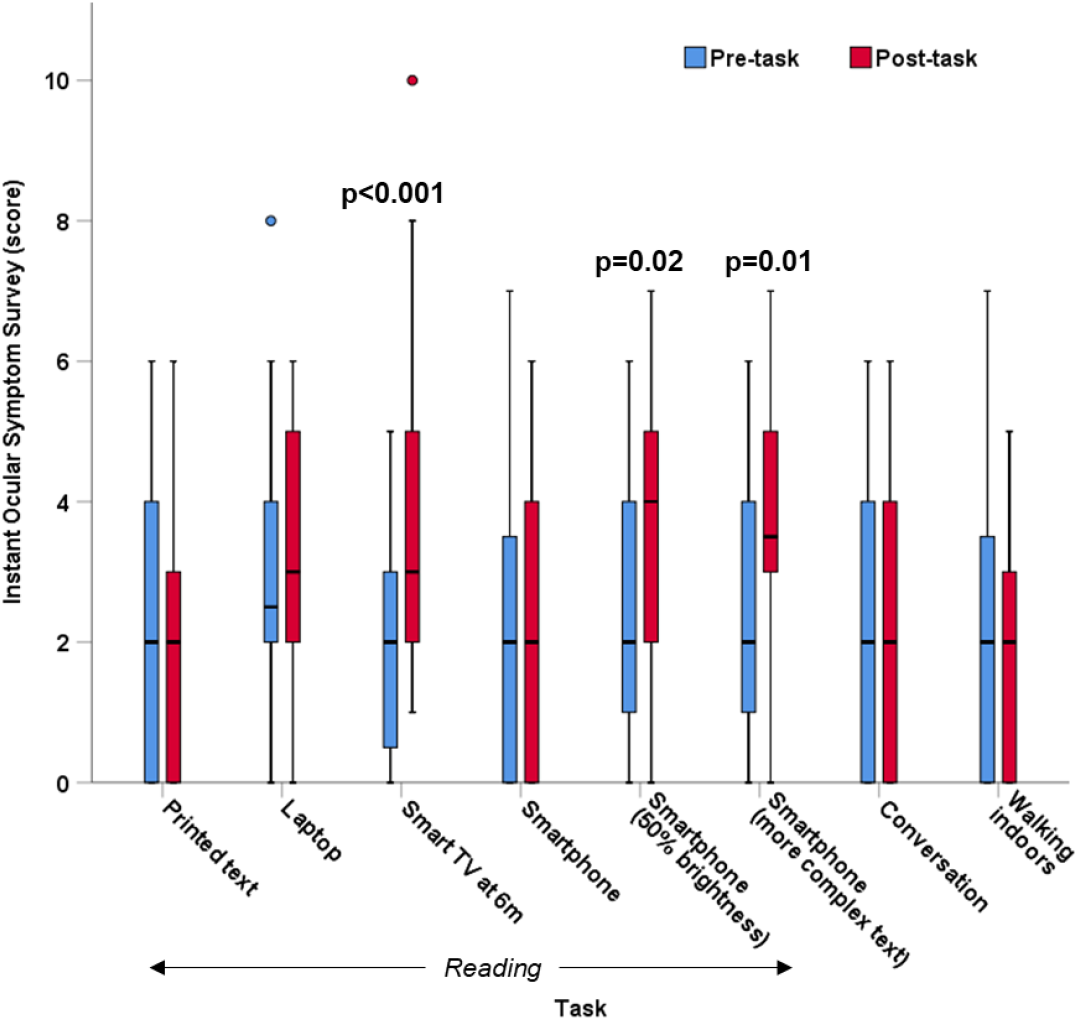
Ocular surface symptoms of discomfort and dryness scores (median and IQR) measured using Instant Ocular Symptoms Survey (IOSS) pre- and post-various tasks of 15 minutes duration for 24 students with healthy eyes. Note, data from the first three minutes of the smartphone task were discarded and the remaining 12 minutes used for analysis. Higher IOSS scores indicate worse discomfort. *Blue and red circles represent mild outliers (symptom scores >1*.*5 to 3 times the interquartile range)*.

### 3.3 Repeatability of blink rate and interblink interval

Group mean blink rate for participants while reading from a smartphone was 10.6±10.4 blinks/min for the first repeat and 11.3±10.4 blinks/min for the second repeat. Interblink interval was 10.3±9.7s and 9.7±11.2s for the first and second repeats respectively. There was no significant difference between two repeated measurements for blink rate (p=0.62) or interblink interval (p=0.55). The Bland and Altman plots for blink rate and interblink interval showing the bias and limits of agreement are presented in Figure 6. The CoR was calculated to be ±12.4 blinks/min for blink rate and ±18.8s for interblink interval.

**Figure 6.**
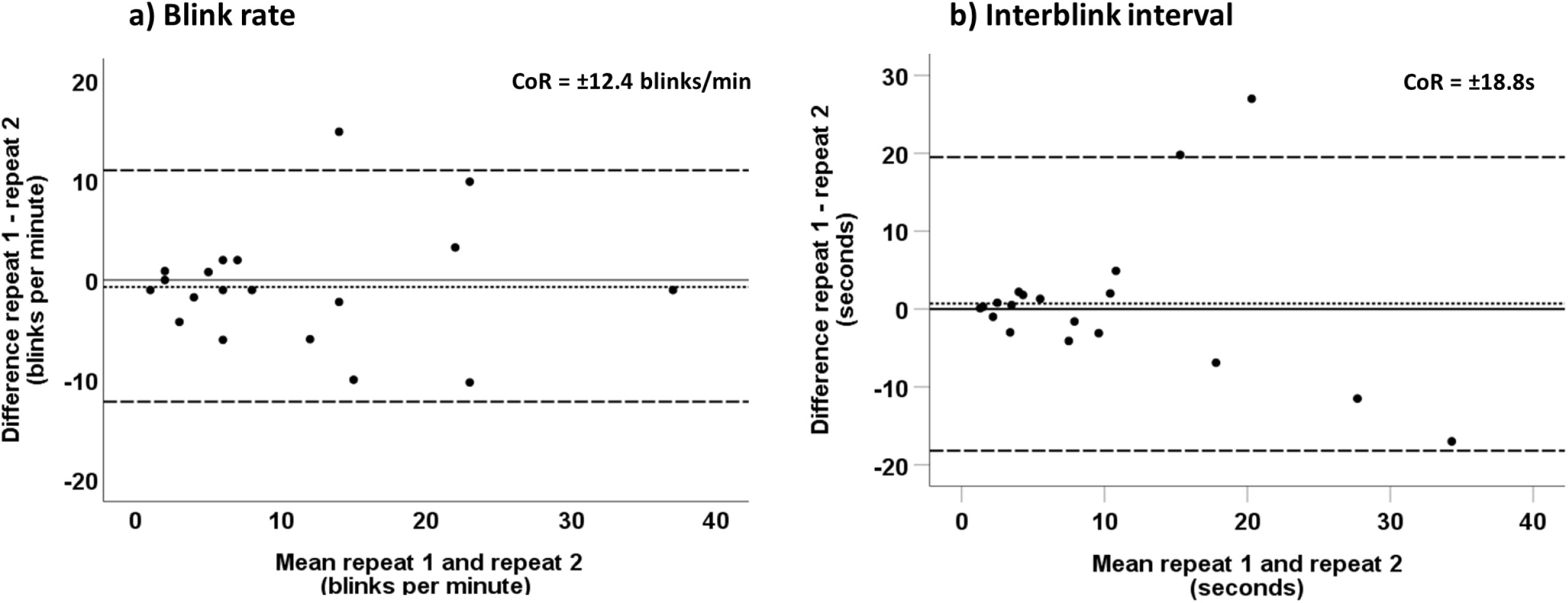
Differences between **a)** blink rate and **b)** interblink interval measured using the wearable eye tracking headset (Pupil Labs GmbH Berlin, Germany) during two repeats plotted against their mean for 24 students with healthy eyes, while reading easy book series on a smartphone for 12 minutes. The dotted line shows a bias of **a)** -0.7 blinks/min (p=0.62) and **b)** 0.7s (p=0.55). The dashed lines represent the limits of agreement of **a)** +11.7 to -13.1 blinks/min and **b)** +19.5 to -18.2s. CoR is the coefficient of repeatability.

## 4. Discussion

A wearable eye tracking headset can be utilised to reliably measure blinking in a variety of real-life settings and was found to be repeatable day to day. Blink rate was consistently reduced and interblink interval was longer during reading compared to conversation or walking, irrespective of reading task complexity, screen brightness, working distance or device used. Changes in blink rate and interblink interval occurred immediately upon starting tasks and did not change throughout the 15-minute duration. No relationship was apparent between blinking and ocular surface comfort or clinical signs.

Blink rate during reading (mean for all reading tasks 10.7±9.7 blinks/min) and conversation (32.4±12.4 blinks/min) in this study aligns with previous findings of a slower blink rate while reading printed text and on a computer (pooled mean 7.9±3.3 blinks/min) than during conversation in adults (mean 21 blinks/min) [4] and in children (20.5 blinks/min) [26]. A reduced blink rate has been consistently reported with computer or smartphone reading and gaming relative to conversation [27, 61, 62] and rest or primary gaze [5, 7, 12-14, 57, 63]. Blink rate while reading on a smartphone (10.6 blinks/min) is similar to a previously reported mean of 8.9 blinks/min within one minute of gaming on a smartphone [27] and median of 12.5 blinks/min within 10 minutes of reading on a smartphone [50]. A reduced blink rate during reading tasks compared to conversation and walking is as expected, as tasks involving higher cognitive demand and concentration are associated with slower blink rate compared to tasks of lower cognitive demand [4, 5, 8, 9, 64, 65].

Interblink interval measured during conversation in this study (1.5±0.6s) is shorter compared to the only previously reported value of 6±3s in healthy adults [66]. Interblink interval has not been previously measured during reading or walking. Other reports were during rest, predetermined gaze, or steady fixation, and viewing a game or movie on computer with reported mean interblink interval values ranging from 3 to 10 seconds [15, 18, 19, 66-69]. As with blink rate, interblink interval has been speculated to be unconsciously adjusted depending on the importance of perceived visual information - prolonged with greater cognitive demand [68].

Enabled by the portability of the wearable eye tracking headset, this was the first study to report blink rate while walking. Blinking during walking did not differ from conversation. A previous study speculated that the cognitive demand during conversation compares to that during orientation simulated in a laboratory, similar to walking [70].

Blinking was not affected by the type of device used in this study. These results align with previous reports that blink rate remains unchanged when an identical reading task is performed in print and on any type of digital device [8, 9, 11, 63, 71, 72].

Text complexity did not modulate the effect of reading on blinking in this study, in agreement with a previous study which compared blink rate while reading regular words with re-ordered mirrored images of the same words [65]. In contrast, another study found a small reduction in blink rate during complex reading compared to non-complex reading on tablet and printed text [8]. Other studies [3, 5, 10, 63] which report reduced blinking while reading complex text on a computer, tablet or printed text did not directly compare texts of differing complexities. The likely high reading comprehension ability level of university student participants may have limited this study’s ability to demonstrate an effect of text complexity.

Screen brightness did not affect blink rate in the present study. Another study found reduced blinking while reading from computer with high screen brightness compared to low screen brightness [73]. The higher blinking with low screen brightness under standard background luminance was speculated to be caused by increased glare discomfort [21, 73]. An effect of screen brightness on blink rate may not be expected as in the photopic range, the eye and visual system constantly and rapidly adapts to luminance changes [74].

The presentation of the reading tasks at near or distance did not impact blinking in this study. A relationship between screen viewing distance and blink rate has not been previously reported. Direction of gaze during tasks may also modulate blink behaviour. Tasks involving down gaze such as reading on printed text and smartphone, may be less likely to trigger blinking compared to tasks involving upward (e.g., smart TV at 6m) [21, 67] or primary gaze [65]. Upward gaze direction could lead to ocular surface area exposure, thereby stimulating blinks [9, 21, 75].

The impact of task on blink rate and interblink interval in this study was immediate and remained unchanged throughout task duration, in agreement with earlier work. A study that investigated blink rate each 30s over the course of 10 minutes reading on a tablet also found no changes in blink rate [8]. A study using the same eye tracker device in school-aged children similarly found a rapid slowing of blink rate and lengthening of interblink interval which occurred within the first minute of gaming on a smartphone, and this remained further unchanged throughout one hour of gaming [27]. A study in adults found no difference in blink rate over the course of one hour gaming on a smartphone [76]. An intervention study in adults found an increase in rate of incomplete blinks from 1 to 60 minutes of smartphone reading, but no change in rate of complete blinks [77].

Ocular symptoms worsened when reading on a smartphone with 50% screen brightness, more complex text on smartphone and reading on smart TV at 6m but there was no association between these changes in symptoms and blink rate or interblink interval. Whereas previous studies in adults did not find direct associations between symptoms and blink rate during digital device use similar to this study [3, 11, 22], increased occurrence of incomplete blinking has been implicated in the worsening of ocular symptoms while reading on a computer or smartphone [11, 22, 77, 78]. Complete blinking is essential to replenishment of the tear film and maintenance of ocular comfort [79] and incomplete blinking can potentially impact dry eye symptoms [80, 81]. Blink amplitude was not characterised in the current study but its usefulness as a possible marker of ocular surface health warrants exploration.

Day to day repeatability of blink rate was CoR ±12.4 blinks/min; this sets the smallest measurable difference in blink rate in longitudinal studies. A closer inspection of the limits of agreement in Figure 6 suggests that CoR may differ with magnitude of blink measurement. Therefore, the CoR was calculated separately for blink rate values higher than 10 blinks/min. Repeatability was better with blink rates ≤ 10 blinks/min (CoR: ±5.4 blinks/min) but less reliable above 10 blinks/min (CoR: ±18.8 blinks/min) (supplementary data Table 3). Previous studies intending to report repeatability of blink rate do not provide a standard repeatability measure to enable comparison with the present findings [6, 18, 28, 29]. These findings suggest that the wearable eye tracker is able to reliably measure blink rate within the normal ranges of spontaneous blinking. These results provide a basis on which to estimate sample size in future studies.

As for blink rate, the repeatability of interblink interval was better for values below 10s (CoR: ±3.9s) and poorer for longer interblink interval values (CoR: ±29.6s) (supplementary data Table 3). The overall CoR for interblink interval is higher compared to the normal range previously reported [19]. No studies have previously examined repeatability of interblink interval.

The strengths of this study lie with measurement of blinking *in situ* without the need for head restraint. Also, blink rate and interblink interval were compared between various tasks on differing devices, controlling for complexity, viewing distance, direction of gaze, and luminance, within the one study. Based on these results, *in situ* measurements of blinking parameters may not be feasible in a small proportion of participants due to poor pupil detection confidence the causes of which require further investigation. Poor pupil confidence unrelated to blinks can occur when using the wearable headset due to extreme gaze angles or pupil obscuration by eyelashes [58]. Excluded data in the current study was likely unrelated to gaze angle, as no extreme gaze angles were observed by the continuous eye monitoring during data collection. Future studies should explore whether this limitation is uniquely related to participant’s eye characteristics (e.g., long eye lashes) [58].

Incomplete blinking has been reported as an important marker of ocular symptoms during reading on smartphone and computer [9, 11, 77] and also during driving [82]. Future studies using the wearable eye tracker will enable examination of blink amplitude *in situ* during various task and conditions.

## 5. Conclusion

Blink rate was reduced and interblink interval increased during reading compared to conversation and walking. Changes in blink rate and interblink interval were immediate and sustained for all tasks, suggesting blinking is a rapidly responsive marker of changes. The similarity in blink rate and interblink interval response during a variety of reading tasks, including smartphone, suggests that reduced blink rate during reading is not driven by type of device used, working distance, screen brightness, nor duration or complexity of task, but rather is intrinsic to the task of reading itself. There was no apparent relationship between changes in blinking and ocular surface comfort or signs.

Blink rate measured using a wearable device *in situ* was repeatable day to day. The current study established solid foundations for the usefulness of blinking as a repeatable and responsive marker of ocular surface health when measured *in situ*. Future research should explore its utility in the settings of dry eye diagnosis and monitoring of treatment effectiveness.

## Data Availability

The datasets generated during and/or analysed during the current study are available in the Mendeley Data repository

https://data.mendeley.com/drafts/j63x6bxj8k

## Acknowledgements

Dr. Peter Wagner for technical support during data collection using the wearable eye tracking headset. The authors acknowledge the Eye Research Group at the School of Optometry and Vision Science for the provision of clinical facilities in support of this research.

## Data Availability Statement

The datasets generated during and/or analysed during the current study are available in the Mendeley Data repository, https://data.mendeley.com/drafts/j63×6bxj8k, Doi: 10.17632/j63×6bxj8k.1

## Notes

**Funding** This research did not receive specific funding from agencies in the public, commercial, or not-for-profit sectors. The first author, NC received a UNSW Tuition Fee Remission Postgraduate Research Scholarship and the Australian Government Research Training Program Thesis Completion Scholarship. The research was also supported by the Dorothy Carlborg Research Grant from the Cornea and Contact Lens Society of Australia, and the UNSW Faculty of Science Research Infrastructure Scheme. The funding sources have no involvement in the study design, conduct of the research, collection, analysis and interpretation of data, writing of the report, preparation of the article or in the decision to submit the article for publication.

### Competing Interest Statement

The authors have declared no competing interest.

### Funding Statement

This research did not receive specific funding from agencies in the public, commercial, or not-for-profit sectors.
The first author, NC received a UNSW Tuition Fee Remission Postgraduate Research Scholarship and the Australian Government Research Training Program Thesis Completion Scholarship. The research was also supported by the Dorothy Carlborg Research Grant from the Cornea and Contact Lens Society of Australia, and the UNSW Faculty of Science Research Infrastructure Scheme. The funding sources have no involvement in the study design, conduct of the research, collection, analysis and interpretation of data, writing of the report, preparation of the article or in the decision to submit the article for publication.

### Author Declarations

Approval was obtained from the UNSW Sydney Human Research Ethics Advisory Panel

