## Supplementary data for "Blink rate measured *in situ* decreases while reading from printed text or digital devices, regardless of task duration, complexity or working distance"

Supplementary **Table 1.** Spontaneous blink parameters (blink rate, interblink interval) measured using a wearable eye tracking headset (Pupil Labs GmbH Berlin, Germany) and associations with change in ocular symptoms measured using Instant Ocular Symptom Survey (IOSS) between tasks for 24 university students with healthy eyes. The p-values were adjusted for multiple comparisons using the one-step Bonferroni method.

| **Change in ocular symptom by task** | **Blink rate (per minute)** | | **Interblink interval (seconds)** | |
|---|---|---|---|---|
| | ***correlation*** | **p-value** | ***correlation*** | **p-value** |
| Printed Text | 0.33 | 1.00 | -0.39 | 1.00 |
| Laptop | 0.06 | 1.00 | 0.11 | 1.00 |
| Smart TV at 6m | 0.25 | 1.00 | -0.26 | 1.00 |
| Smartphone | 0.09 | 1.00 | -0.09 | 1.00 |
| Smartphone (50% brightness) | -0.33 | 1.00 | 0.41 | 1.00 |
| Smartphone (more complex text) | -0.10 | 1.00 | 0.05 | 1.00 |
| Conversation | 0.01 | 1.00 | 0.08 | 1.00 |
| Walking indoors | 0.16 | 1.00 | -0.09 | 1.00 |

Supplementary **Table 2.** Associations between blink parameters (blink rate, interblink interval) measured using the Pupil Labs wearable eye tracking headset with Pupil software v2.0 during various reading and non-reading tasks, and ocular surface symptoms, tear film function and meibomian gland characteristics for 24 university students with healthy eyes. The p-values were adjusted for multiple comparisons using the one-step Bonferroni method. *Abbreviations: IOSS, Instant Ocular Symptoms Survey; DEQ-5, Dry Eye Questionnaire 5; SANDE, Symptoms Assessment in Dry Eye; OSDI, Ocular Surface Disease Index.*

| **Variables** | **Blink rate (per minute)** | | **Interblink interval (seconds)** | |
|---|---|---|---|---|
| | ***correlation*** | **p-value** | ***correlation*** | **p-value** |
| **Printed text** | | | | |
| ***Ocular surface symptoms* (score)** | | | | |
| IOSS | -0.01 | 1.00 | 0.04 | 1.00 |
| DEQ-5 | 0.07 | 1.00 | -0.01 | 1.00 |
| SANDE | -0.16 | 1.00 | 0.22 | 1.00 |
| OSDI | 0.29 | 1.00 | -0.26 | 1.00 |
| ***Tear film function*** | | | | |
| Lipid layer thickness (nm) | -0.01 | 1.00 | 0.07 | 1.00 |
| Tear meniscus height (mm) | 0.30 | 1.00 | -0.32 | 1.00 |
| Non-invasive tear break-up time (s) | -0.23 | 1.00 | 0.25 | 1.00 |
| ***Meibomian gland*** | | | | |
| Eyelid telangiectasia (grade) | -0.05 | 1.00 | 0.08 | 1.00 |
| Expressibility (number of expressible glands) | -0.10 | 1.00 | -0.10 | 1.00 |
| Expressibility (amount of pressure applied) | -0.09 | 1.00 | -0.14 | 1.00 |
| Expressed meibum quality (grade) | 0.13 | 1.00 | -0.18 | 1.00 |
| ***Meibography*** | | | | |
| Meibomian gland area loss (score) | 0.11 | 1.00 | -0.00 | 1.00 |
| Meibomian gland morphological pattern present: | | | | |
| Dilation | 0.14 | 1.00 | -0.08 | 1.00 |
| Shortening | 0.22 | 1.00 | -0.29 | 1.00 |
| Tortuosity | -016 | 1.00 | 0.13 | 1.00 |
| **Laptop** | | | | |
| ***Ocular surface symptoms* (score)** | | | | |
| IOSS | -0.08 | 1.00 | 0.08 | 1.00 |
| DEQ-5 | -0.06 | 1.00 | 0.02 | 1.00 |
| SANDE | -0.19 | 1.00 | 0.17 | 1.00 |
| OSDI | 0.01 | 1.00 | -0.20 | 1.00 |
| ***Tear film function*** | | | | |
| Lipid layer thickness (nm) | -0.04 | 1.00 | -0.00 | 1.00 |
| Tear meniscus height (mm) | 0.42 | 0.60 | -0.49 | 0.30 |
| Non-invasive tear break-up time (s) | -0.06 | 1.00 | 0.06 | 1.00 |
| ***Meibomian gland*** | | | | |
| Eyelid telangiectasia (grade) | -0.02 | 1.00 | 0.09 | 1.00 |
| Expressibility (number of expressible glands) | 0.18 | 1.00 | -0.14 | 1.00 |
| Expressibility (amount of pressure applied) | 0.24 | 1.00 | -0.22 | 1.00 |
| Expressed meibum quality (grade) | 0.31 | 1.00 | -0.19 | 1.00 |
| ***Meibography*** | | | | |
| Meibomian gland area loss (score) | -0.25 | 1.00 | 0.18 | 1.00 |
| Meibomian gland morphological pattern present: | | | | |
| Dilation | -0.09 | 1.00 | 0.07 | 1.00 |

| | | | | |
|---|---|---|---|---|
| Shortening | 0.09 | 1.00 | -0.19 | 1.00 |
| Tortuosity | -0.29 | 1.00 | 0.23 | 1.00 |
| **Smart TV at 6m** | | | | |
| ***Ocular surface symptoms* (score)** | | | | |
| IOSS | 0.26 | 1.00 | -0.25 | 1.00 |
| DEQ-5 | 0.12 | 1.00 | 0.10 | 1.00 |
| SANDE | 0.19 | 1.00 | -0.19 | 1.00 |
| OSDI | 0.42 | 0.75 | -0.40 | 0.90 |
| ***Tear film function*** | | | | |
| Lipid layer thickness (nm) | -0.70 | 1.00 | 0.11 | 1.00 |
| Tear meniscus height (mm) | 0.34 | 1.00 | -0.34 | 1.00 |
| Non-invasive tear break-up time (s) | -0.04 | 1.00 | 0.06 | 1.00 |
| ***Meibomian gland*** | | | | |
| Eyelid telangiectasia (grade) | -0.14 | 1.00 | 0.10 | 1.00 |
| Expressibility (number of expressible glands) | -0.13 | 1.00 | 0.14 | 1.00 |
| Expressibility (amount of pressure applied) | -0.24 | 1.00 | 0.24 | 1.00 |
| Expressed meibum quality (grade) | 0.14 | 1.00 | -0.14 | 1.00 |
| ***Meibography*** | | | | |
| Meibomian gland area loss (score) | -0.33 | 1.00 | 0.31 | 1.00 |
| Meibomian gland morphological pattern present: | | | | |
| Dilation | -0.08 | 1.00 | 0.07 | 1.00 |
| Shortening | 0.14 | 1.00 | -0.18 | 1.00 |
| Tortuosity | -012 | 1.00 | 0.09 | 1.00 |
| **Smartphone** | | | | |
| ***Ocular surface symptoms* (score)** | | | | |
| IOSS | 0.03 | 1.00 | 0.01 | 1.00 |
| DEQ-5 | 0.15 | 1.00 | -0.15 | 1.00 |
| SANDE | -0.01 | 1.00 | 0.03 | 1.00 |
| OSDI | 0.19 | 1.00 | -0.11 | 1.00 |
| ***Tear film function*** | | | | |
| Lipid layer thickness (nm) | 0.09 | 1.00 | -0.07 | 1.00 |
| Tear meniscus height (mm) | 0.24 | 1.00 | -0.29 | 1.00 |
| Non-invasive tear break-up time (s) | -0.44 | 0.90 | 0.45 | 0.75 |
| ***Meibomian gland*** | | | | |
| Eyelid telangiectasia (grade) | 0.03 | 1.00 | 0.00 | 1.00 |
| Expressibility (number of expressible glands) | 0.07 | 1.00 | -0.12 | 1.00 |
| Expressibility (amount of pressure applied) | -0.14 | 1.00 | 0.06 | 1.00 |
| Expressed meibum quality (grade) | -0.15 | 1.00 | 0.12 | 1.00 |
| ***Meibography*** | | | | |
| Meibomian gland area loss (score) | -0.01 | 1.00 | 0.01 | 1.00 |
| Meibomian gland morphological pattern present: | | | | |
| Dilation | 0.45 | 0.75 | -0.40 | 1.00 |
| Shortening | 0.36 | 1.00 | -0.39 | 1.00 |
| Tortuosity | -0.21 | 1.00 | 0.21 | 1.00 |
| **Smartphone (50% brightness)** | | | | |

| | | | | |
|---|---|---|---|---|
| ***Ocular surface symptoms* (score)** | | | | |
| IOSS | -0.06 | 1.00 | -0.02 | 1.00 |
| DEQ-5 | 0.04 | 1.00 | 0.02 | 1.00 |
| SANDE | -0.14 | 1.00 | 0.18 | 1.00 |
| OSDI | 0.09 | 1.00 | -0.05 | 1.00 |
| ***Tear film function*** | | | | |
| Lipid layer thickness (nm) | -0.02 | 1.00 | 0.15 | 1.00 |
| Tear meniscus height (mm) | 0.24 | 1.00 | -0.19 | 1.00 |
| Non-invasive tear break-up time (s) | -0.32 | 1.00 | 0.46 | 0.45 |
| ***Meibomian gland*** | | | | |
| Eyelid telangiectasia (grade) | 0.38 | 1.00 | 0.02 | 1.00 |
| Expressibility (number of expressible glands) | 0.11 | 1.00 | -0.27 | 1.00 |
| Expressibility (amount of pressure applied) | -0.07 | 1.00 | -0.06 | 1.00 |
| Expressed meibum quality (grade) | -0.06 | 1.00 | -0.02 | 1.00 |
| ***Meibography*** | | | | |
| Meibomian gland area loss (score) | -0.12 | 1.00 | 0.09 | 1.00 |
| Meibomian gland morphological pattern present: | | | | |
| Dilation | 0.20 | 1.00 | -0.12 | 1.00 |
| Shortening | 0.32 | 1.00 | -0.38 | 0.98 |
| Tortuosity | -0.19 | 1.00 | 0.05 | 1.00 |
| **Smartphone (more complex text)** | | | | |
| ***Ocular surface symptoms* (score)** | | | | |
| IOSS | 0.03 | 1.00 | -0.04 | 1.00 |
| DEQ-5 | 0.15 | 1.00 | -0.06 | 1.00 |
| SANDE | 0.02 | 1.00 | 0.05 | 1.00 |
| OSDI | 0.39 | 1.00 | -0.29 | 1.00 |
| ***Tear film function*** | | | | |
| Lipid layer thickness (nm) | 0.19 | 1.00 | -0.12 | 1.00 |
| Tear meniscus height (mm) | 0.33 | 1.00 | -0.45 | 0.60 |
| Non-invasive tear break-up time (s) | -0.27 | 1.00 | 0.32 | 1.00 |
| ***Meibomian gland*** | | | | |
| Eyelid telangiectasia (grade) | -0.27 | 1.00 | 0.23 | 1.00 |
| Expressibility (number of expressible glands) | 0.09 | 1.00 | -0.13 | 1.00 |
| Expressibility (amount of pressure applied) | 0.03 | 1.00 | -0.05 | 1.00 |
| Expressed meibum quality (grade) | 0.16 | 1.00 | -0.25 | 1.00 |
| ***Meibography*** | | | | |
| Meibomian gland area loss (score) | -0.29 | 1.00 | 0.25 | 1.00 |
| Meibomian gland morphological pattern present: | | | | |
| Dilation | 0.20 | 1.00 | -0.15 | 1.00 |
| Shortening | 0.07 | 1.00 | -0.09 | 1.00 |
| Tortuosity | -0.17 | 1.00 | 0.14 | 1.00 |
| **Conversation** | | | | |
| ***Ocular surface symptoms* (score)** | | | | |
| IOSS | -0.14 | 1.00 | 0.16 | 1.00 |
| DEQ-5 | -0.16 | 1.00 | 0.16 | 1.00 |
| SANDE | -0.16 | 1.00 | 0.23 | 1.00 |

| | | | | |
|---|---|---|---|---|
| OSDI | 0.04 | 1.00 | 0.06 | 1.00 |
| ***Tear film function*** | | | | |
| Lipid layer thickness (nm) | -0.09 | 1.00 | 0.09 | 1.00 |
| Tear meniscus height (mm) | 0.33 | 1.00 | -0.29 | 1.00 |
| Non-invasive tear break-up time (s) | -0.33 | 1.00 | 0.42 | 0.75 |
| ***Meibomian gland*** | | | | |
| Eyelid telangiectasia (grade) | 0.06 | 1.00 | -0.18 | 1.00 |
| Expressibility (number of expressible glands) | 0.02 | 1.00 | -0.06 | 1.00 |
| Expressibility (amount of pressure applied) | 0.03 | 1.00 | -0.03 | 1.00 |
| Expressed meibum quality (grade) | 0.56 | 0.15 | -0.43 | 0.98 |
| ***Meibography*** | | | | |
| Meibomian gland area loss (score) | -0.18 | 1.00 | 0.22 | 1.00 |
| Meibomian gland morphological pattern present: | | | | |
| Dilation | -0.04 | 1.00 | 0.00 | 1.00 |
| Shortening | 0.09 | 1.00 | -0.18 | 1.00 |
| Tortuosity | -0.02 | 1.00 | -0.09 | 1.00 |
| **Walking indoors** | | | | |
| ***Ocular surface symptoms* (score)** | | | | |
| IOSS | -0.10 | 1.00 | 0.14 | 1.00 |
| DEQ-5 | -0.10 | 1.00 | 0.09 | 1.00 |
| SANDE | -0.03 | 1.00 | 0.03 | 1.00 |
| OSDI | 0.23 | 1.00 | -0.21 | 1.00 |
| ***Tear film function*** | | | | |
| Lipid layer thickness (nm) | -0.26 | 1.00 | 0.25 | 1.00 |
| Tear meniscus height (mm) | 0.18 | 1.00 | -0.19 | 1.00 |
| Non-invasive tear break-up time (s) | -0.20 | 1.00 | 0.18 | 1.00 |
| ***Meibomian gland*** | | | | |
| Eyelid telangiectasia (grade) | 0.18 | 1.00 | -0.14 | 1.00 |
| Expressibility (number of expressible glands) | -0.20 | 1.00 | 0.30 | 1.00 |
| Expressibility (amount of pressure applied) | -0.19 | 1.00 | 0.26 | 1.00 |
| Expressed meibum quality (grade) | 0.39 | 1.00 | -0.31 | 1.00 |
| ***Meibography*** | | | | |
| Meibomian gland area loss (score) | 0.06 | 1.00 | 0.01 | 1.00 |
| Meibomian gland morphological pattern present: | | | | |
| Dilation | -0.03 | 1.00 | -0.03 | 1.00 |
| Shortening | 0.11 | 1.00 | -0.01 | 1.00 |
| Tortuosity | -0.15 | 1.00 | 0.16 | 1.00 |

Supplementary **Table 3.** The mean differences, limits of agreement and coefficient of repeatability of blink rate and interblink interval values ≤ 10 and > 10, measured using the wearable eye tracking headset (Pupil Labs GmbH Berlin, Germany) during two repeats for 24 university students with healthy eyes while reading on a smartphone for 12 minutes.

| **Repeatability** | **Blink rate (blinks/min): values ≤ 10** | **Blink rate (blinks/min): values > 10** | **Interblink interval (s): values ≤ 10** | **Interblink interval (s): values > 10** |
|---|---|---|---|---|
| Mean first repeat | 3.8 | 19.9 | 3.8 | 19.2 |
| Mean second repeat | 4.7 | 20.4 | 4.1 | 17.3 |
| Mean difference (bias) between repeats (p-value) | -0.9 (p = 0.30) | -0.5 (p = 0.89) | -0.2 (p = 0.69) | 1.9 (p = 0.73) |
| Limits of agreement (LOA = bias±CoR) | +4.5 to -6.3 | +18.3 to -19.3 | +3.7 to -4.2 | +31.5 to -27.7 |
| Coefficient of repeatability (CoR = 1.96 X SD of differences between the two repeats) | ±5.4 | ±18.8 | ±3.9 | ±29.6 |
